# Five dominant dimensions of brain aging are identified via deep learning: associations with clinical, lifestyle, and genetic measures

**DOI:** 10.1101/2023.12.29.23300642

**Authors:** Zhijian Yang, Junhao Wen, Guray Erus, Sindhuja T. Govindarajan, Randa Melhem, Elizabeth Mamourian, Yuhan Cui, Dhivya Srinivasan, Ahmed Abdulkadir, Paraskevi Parmpi, Katharina Wittfeld, Hans J. Grabe, Robin Bülow, Stefan Frenzel, Duygu Tosun, Murat Bilgel, Yang An, Dahyun Yi, Daniel S. Marcus, Pamela LaMontagne, Tammie L.S. Benzinger, Susan R. Heckbert, Thomas R. Austin, Shari R. Waldstein, Michele K. Evans, Alan B. Zonderman, Lenore J. Launer, Aristeidis Sotiras, Mark A. Espeland, Colin L. Masters, Paul Maruff, Jurgen Fripp, Arthur Toga, Sid O’Bryant, Mallar M. Chakravarty, Sylvia Villeneuve, Sterling C. Johnson, John C. Morris, Marilyn S. Albert, Kristine Yaffe, Henry Völzke, Luigi Ferrucci, Nick R. Bryan, Russell T. Shinohara, Yong Fan, Mohamad Habes, Paris Alexandros Lalousis, Nikolaos Koutsouleris, David A. Wolk, Susan M. Resnick, Haochang Shou, Ilya M. Nasrallah, Christos Davatzikos

**Affiliations:** Artificial Intelligence in Biomedical Imaging Laboratory (AIBIL), Center for and Data Science for Integrated Diagnostics (AI2D), Perelman School of Medicine, University of Pennsylvania, Philadelphia, PA, USA; Graduate Group in Applied Mathematics and Computational Science, University of Pennsylvania, Philadelphia, PA, USA; Laboratory of AI and Biomedical Science (LABS), Stevens Neuroimaging and Informatics Institute, Keck School of Medicine of USC, University of Southern California, Los Angeles, California, USA; Laboratory for Research in Neuroimaging, Department of Clinical Neurosciences, Lausanne University Hospital (CHUV) and University of Lausanne, Lausanne, Switzerland; Department of Psychiatry and Psychotherapy, University Medicine Greifswald, Germany; German Center for Neurodegenerative Diseases (DZNE), Site Rostock/Greifswald, Germany; Institute of Diagnostic Radiology and Neuroradiology, University of Greifswald, Germany; Department of Radiology and Biomedical Imaging, University of California, San Francisco, San Francisco, CA, USA; Laboratory of Behavioral Neuroscience, National Institute on Aging, National Institutes of Health, Baltimore, MD, USA; Institute of Human Behavioral Medicine, Medical Research Center Seoul National University, Seoul, Republic of Korea; Mallinckrodt Institute of Radiology, Washington University School of Medicine, St. Louis, MO, USA; Cardiovascular Health Research Unit and Department of Epidemiology, University of Washington, Seattle, WA, USA; Department of Psychology, University of Maryland, Baltimore County, Catonsville, MD, USA; Health Disparities Research Section, Laboratory of Epidemiology and Population Sciences, NIA/NIH/IRP, Baltimore, MD, USA; Neuroepidemiology Section, Intramural Research Program, National Institute on Aging, Bethesda, Maryland, USA; Department of Radiology and Institute of Informatics, Washington University in St. Luis, St. Luis, MO63110, USA; Department of Internal Medicine, Wake Forest University School of Medicine, Winston-Salem, NC, USA; Florey Institute, The University of Melbourne, Parkville, VIC, 3052, Australia; CSIRO Health and Biosecurity, Australian e-Health Research Centre CSIRO, Brisbane, Queensland, Australia; Laboratory of Neuro Imaging, USC Stevens Neuroimaging and Informatics Institute, Keck School of Medicine of USC, University of Southern California, Los Angeles, California, USA; Institute for Translational Research University of North Texas Health Science Center Fort Worth Texas USA.; Computational Brain Anatomy (CoBrA) Laboratory, Cerebral Imaging Center, Douglas Mental Health University Institute, McGill University, Verdun, Quebec, Canada; McConnell Brain Imaging Centre, Montreal Neurological Institute, McGill University, Montreal, Canada; Wisconsin Alzheimer’s Institute, University of Wisconsin School of Medicine and Public Health, Madison, Wisconsin, USA; Knight Alzheimer Disease Research Center, Washington University in St. Louis, St. Louis, MO, USA; Department of Neurology, Johns Hopkins University School of Medicine, Baltimore, MD, USA; Departments of Neurology, Psychiatry and Epidemiology and Biostatistics, University of California San Francisco, San Francisco, CA, USA; Institute for Community Medicine, University Medicine Greifswald, Greifswald, Germany; Translational Gerontology Branch, Longitudinal Studies Section, National Institute on Aging, National Institutes of Health, MedStar Harbor Hospital, 3001 S. Hanover Street, Baltimore, MD, USA; Department of Radiology, University of Pennsylvania, Philadelphia, PA, USA; Penn Statistics in Imaging and Visualization Center, Department of Biostatistics, Epidemiology, & Informatics, University of Pennsylvania, Philadelphia, PA, USA; Biggs Alzheimer’s Institute, University of Texas San Antonio Health Science Center, USA; Department of Psychosis Studies, Institute of Psychiatry, Psychology & Neuroscience, King’s College London, London, United Kingdom; Section for Precision Psychiatry, Department of Psychiatry and Psychotherapy, Ludwig-Maximilian-University Munich, Munich, Germany; Department of Neurology, University of Pennsylvania, Philadelphia, PA, USA

**Author notes:** Corresponding author: Christos Davatzikos, PhD – 3700 Hamilton Walk, 7^th^ Floor, Philadelphia, PA 19104.

## Abstract

Brain aging is a complex process influenced by various lifestyle, environmental, and genetic factors, as well as by age-related and often co-existing pathologies. MRI and, more recently, AI methods have been instrumental in understanding the neuroanatomical changes that occur during aging in large and diverse populations. However, the multiplicity and mutual overlap of both pathologic processes and affected brain regions make it difficult to precisely characterize the underlying neurodegenerative profile of an individual from an MRI scan. Herein, we leverage a state-of-the art deep representation learning method, Surreal-GAN, and present both methodological advances and extensive experimental results that allow us to elucidate the heterogeneity of brain aging in a large and diverse cohort of 49,482 individuals from 11 studies. Five dominant patterns of neurodegeneration were identified and quantified for each individual by their respective (herein referred to as) R-indices. Significant associations between R-indices and distinct biomedical, lifestyle, and genetic factors provide insights into the etiology of observed variances. Furthermore, baseline R-indices showed predictive value for disease progression and mortality. These five R-indices contribute to MRI-based precision diagnostics, prognostication, and may inform stratification into clinical trials.

## Introduction

The human brain undergoes structural changes during the aging process, but the trajectories of these changes vary significantly among individuals, highlighting the heterogeneity in brain aging. Various factors, including genetic and lifestyle factors and diseases, contribute to this heterogeneity, either exacerbating or protecting against age-related neuropathological processes^1^. Moreover, relatively subtle brain changes in specific regions or spatial patterns can emerge early in the pre-clinical stages of neurodegenerative diseases such as Alzheimer’s disease^2,3^. Therefore, unraveling the neuroanatomical heterogeneity in brain aging sheds light on the progression of underlying neuropathologic processes and might offer very early diagnostic, prognostic, vulnerability, or resilience markers.

Neuroimaging plays a pivotal role in studying the human brain, enabling direct quantification of these changes *in vivo*^4^ and at a large scale. This has enriched our comprehension of how aging and diseases influence brain structure and function. The majority of previous studies have relied on case-control group comparisons, which are not designed to address heterogeneity across individuals and pathologies. Some machine learning methods, leveraging binary classifications, tried to derive neuroimaging biomarkers of brain aging at the individual level^5,6^. However, these studies still overlook the underlying heterogeneity and derive biomarkers of a typical or averaged pattern of diverse neuroanatomical changes. Various clustering approaches have been deployed to parse the heterogeneity in aging-related neurological diseases from neuroimaging data^7–9^. However, they are usually confounded by numerous variations in brain structure which are not related to aging and neuropathology. Moreover, they aim to cluster each individual into a single subtype, thus overlooking that an individual might have a mixture of underlying pathologies at different stages.

In contrast to previous methods, Surreal-GAN^10^, a recent weakly-supervised, deep representation learning method, offers a novel and general approach applicable to disentangling the heterogeneity of brain aging. By learning multiple *transformations* from a reference (REF) group (e.g., young and healthy individuals) to a target (TAR) group (e.g., older adults or patients with a specific clinical phenotype), the model captures heterogeneous brain changes relative to the reference population and effectively distills them down to low-dimensional representation indices. These indices, herein called R-indices, indicate the severity of individualized brain changes along multiple dimensions, potentially reflecting the stage of a mixture of underlying neuropathological processes that induce deviations from the distribution of a reference brain structure.

The contribution of the current study is twofold. First, we substantially extended the Surreal-GAN methodology by introducing a correlation structure among the R-indices in a reduced representation latent space, thereby capturing interactions among multiple underlying neuropathological processes. Second, we applied this new methodology to a large and harmonized multi-study, multi-site dataset from the iSTAGING consortium^11^, consisting of more than 50,000 participants from 11 neuroimaging studies. As our goal was to capture patterns of brain aging, in our experiments we defined the REF group as participants younger than 50 years old. All other individuals above 50 years old, including those with mild cognitive impairment (MCI) or dementia, were grouped as the TAR group. We therefore established a mathematically principled representation of the dominant dimensions of neuroanatomical brain aging in this cohort, and associated these dimensions with cognitive, clinical, lifestyle, and genetic measures. Additionally, survival analyses demonstrated that brain change along these dimensions predicted future disease progression and mortality.

### Surreal-GAN disentangles heterogeneity of brain aging via dimensional latent representation

Surreal-GAN^10^ utilizes a deep generative model along with a series of effective regularization constraints^10^ to learn one-to-many transformations from brain measurements of a reference (REF) population, such as a pre-aging or healthy control cohort, to a target (TAR) population, such as aging or disease-related cohorts (**Method 1**). Surreal-GAN therefore captures dominant dimensions, or patterns, of brain change related to a condition of interest while minimizing confounding variations. The expressions of brain changes along each of these dimensions are called R-indices. The same participant can have non-exclusive co-expression of different patterns, indicating that multiple pathologic processes are potentially active (**Figure 1a**).

**Figure 1.**
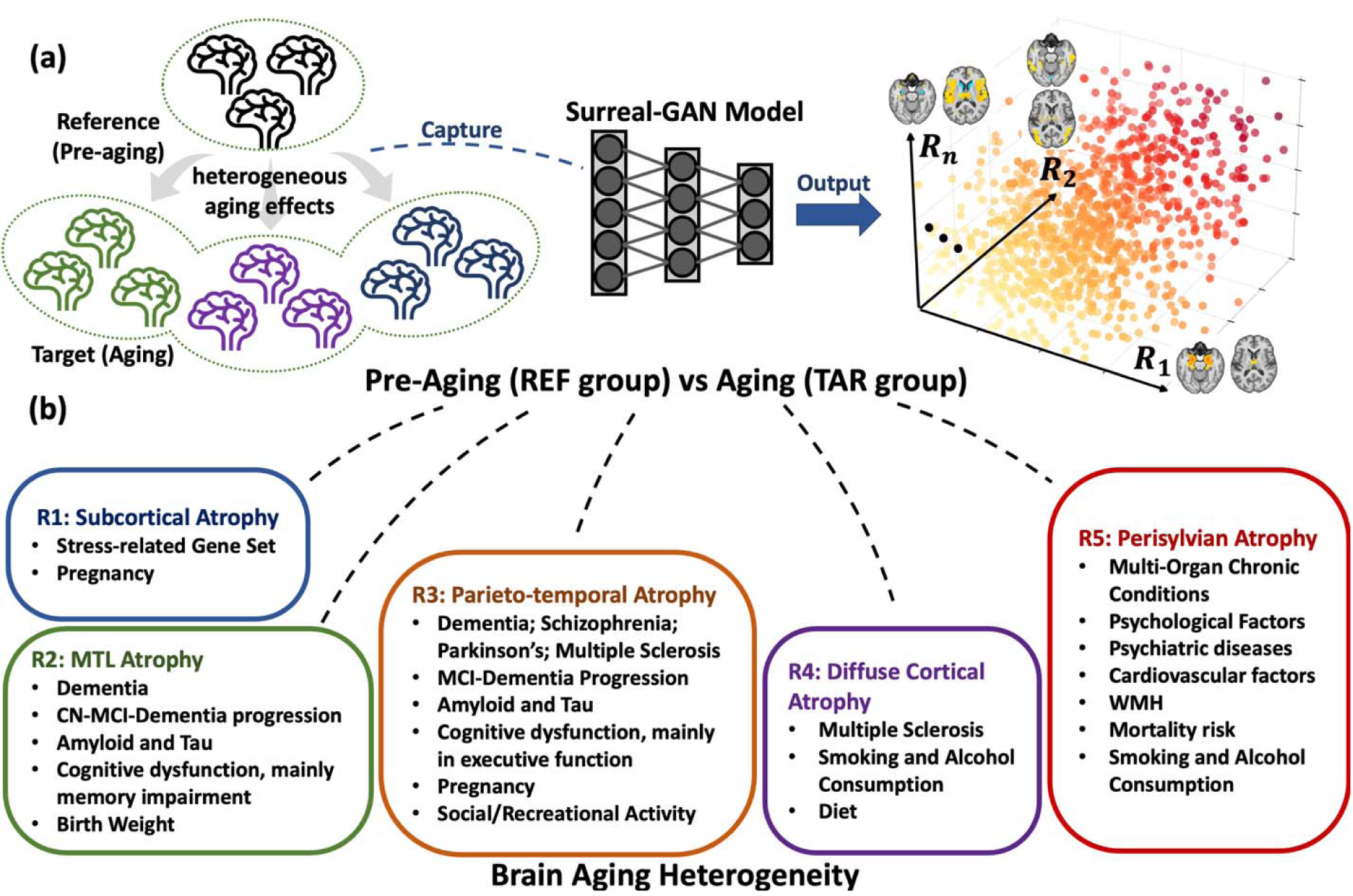
Surreal-GAN disentangles brain aging heterogeneity through a dimensional representation approach. a. The heterogeneous aging effects contribute to distinct alterations in human brain structures, leading to various brain change patterns. Surreal-GAN, an advanced deep learning approach utilizing generative learning, unravels brain variations attributed to the aging process by capturing transformations from a reference (REF) population to a target (TAR) population. It specifically represents the diversity of such brain change patterns using multi-dimensional R-indices. These R-indices serve as indicators of and quantify the type and severity of distinct brain change patterns, which reflect underlying neuropathological processes and their stages. **b.** In this study, to disentangle the neuroanatomical heterogeneity related to brain aging, we set the REF and TAR groups to pre-aging individuals (<50 years old) and all older adults (>50 years old), respectively. Surreal-GAN identifies five reproducible dimensions, each associated with distinct brain change patterns. Further statistical analyses uncover a range of influential factors associated with each dimension, encompassing pathological influences, lifestyle factors, life events, and genetic variants. MTL: medial temporal lobe; CN: cognitively normal; MCI: mild cognitive impairment; WMH: white matter hyperintensities, typically associated with small vessel vascular pathology.

### Improvement of Surreal-GAN by imposing a correlation structure in its latent space

The fundamental framework of the original Surreal-GAN model inherently encourages independence among the derived R-indices. As demonstrated theoretically (**Supplementary eMethod 1.2**), this poses a significant limitation when dealing with correlated ground-truth dimensions or patterns of brain changes. Empirically, we showcased the decline in the model’s performance on semi-synthetic data with different levels of simulated associations among ground-truth patterns. This constraint becomes particularly relevant in the context of brain aging due to the common co-occurrence of multiple underlying pathologies and their potential impact on various brain regions.

In the current work, we addressed this limitation by introducing a parameterization of the correlations among the R-indices using a Gaussian copula during the training process (**Supplementary eMethod 1.2**). The enhanced Surreal-GAN was found to be robust in handling various levels of simulated correlations in semi-synthetic experiments (**Supplementary eResult 1** and **eFigure 1**). In real-data experiments, it effectively derived correlated R-indices, which capture the interactions among underlying pathologic mechanisms (**Figure 2c)**.

**Figure 2.**
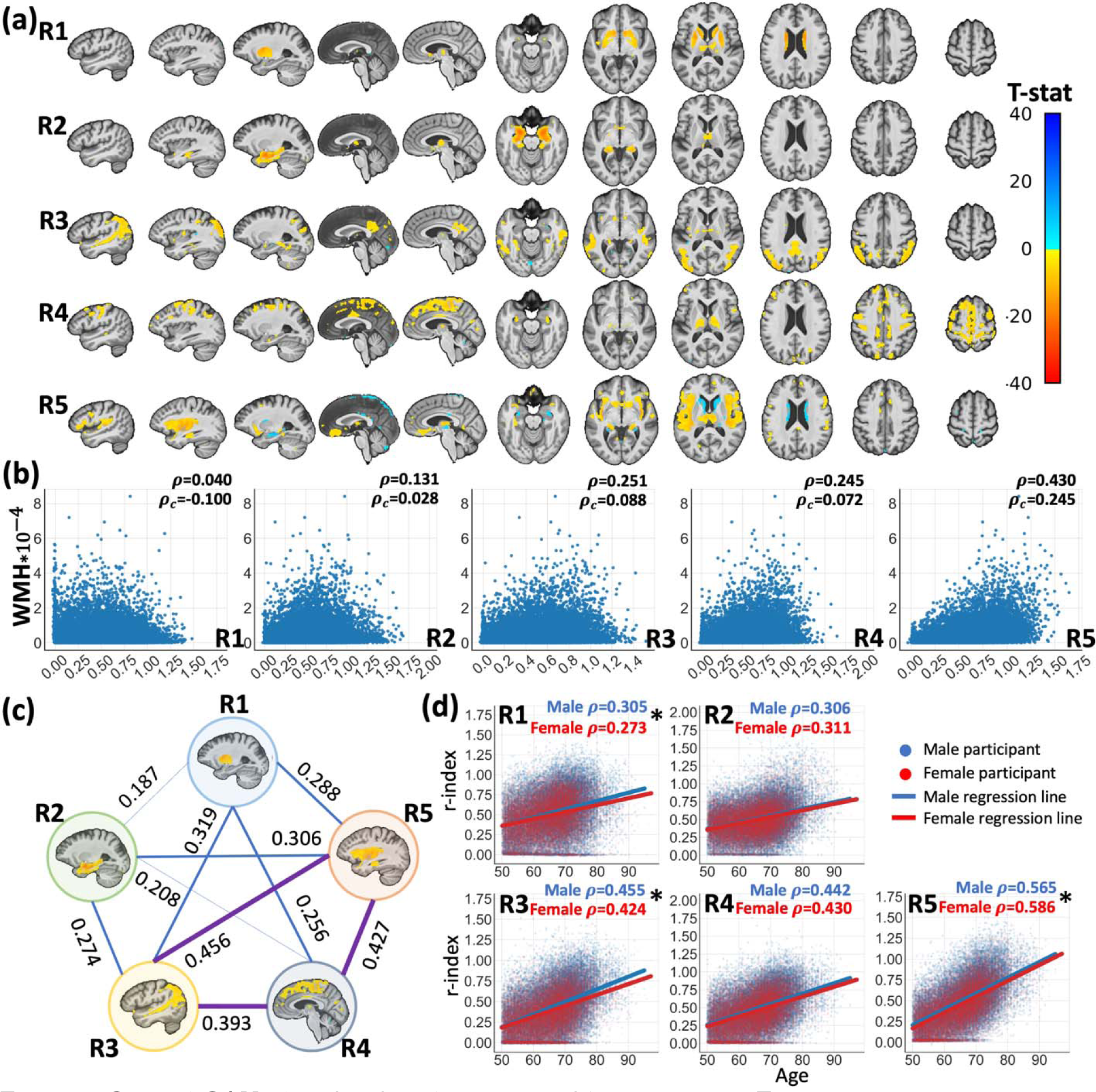
Surreal-GAN identifies five dimensions of brain aging. a. The severity of brain aging along five dimensions in each participant was quantified by the five R-indices (R1-R5), which revealed distinct patterns of associated gray matter atrophy. Characteristic patterns for each R-index are shown via voxel-wise t-tests performed for each R-index while adjusting for age, sex, intracranial volume (ICV), and the remaining four R-indices. False discovery rate (FDR) correction was performed to adjust multiple comparisons with a p-value threshold of 0.001. **b.** The five R-indices show different levels of associations with white matter hyperintensity (WMH) volumes. p_C_ and p denotes associations with and without adjusting for age and sex, respectively. R5 shows the strongest positive associations. **c.** The five R-indices demonstrate positive Pearson correlations with each other, with the strongest associations observed among R3, R4, and R5 **d.** The five R-indices exhibit significant positive associations with chronological age. Additionally, significant differences, as marked by asterisks, were found between males and females in the correlations (p) between age and R1, R3, and R5.

### Application of Surreal-GAN to unravel brain aging heterogeneity

We defined the training set by including a REF group of 1150 participants aged < 50 (i.e., pre- aging participants) and a TAR group of 8992 participants of ages 50-97 including those with MCI or dementia (i.e., all older adults) from 11 studies, thus training the Surreal-GAN model to derive dimensional representations that capture the spectrum of brain aging patterns. Subsequently, we applied the resultant model to all 49,482 iSTAGING participants of ages 50-97 to associate the expression of brain change along each dimension with demographic, clinical, neurocognitive, lifestyle, and genetic measures (**Figure 1b**). Consistency among independently trained models (**Method 3**) suggested that the most reproducible R-indices were derived from five dimensions, which were also replicated using an independent training set consisting of 1000 REF and 4818 TAR participants (**Supplementary eResult 2 and eFigure 2).**

### The five R-indices indicate the severity of distinct but correlated atrophy patterns

We first investigated neuroanatomical changes related to each index using voxel-based morphometry analyses. R1 exhibited significant associations with **subcortical atrophy**, mainly concentrated in the caudate and putamen. R2 was characterized by **focal medial temporal lobe (MTL) atrophy**. R3 indicated the severity of **parieto-temporal atrophy**, including that in middle temporal gyrus, angular gyrus, and middle occipital gyrus. R4 was distinguished by **diffuse cortical atrophy** in medial and lateral frontal regions, as well as superior parietal and occipital regions; R5 primarily indicated **perisylvian atrophy** centered around the insular cortex (**Figure 2a**).

Additionally, we observed significant positive correlations between white matter hyperintensity volumes (WMH) and R2-R5 (**Figure 2b**). Among them, R5 exhibited a much stronger association than other dimensions with and without adjusting for age and sex (p=0.430 and p =0.245, p<1*10^-200^) Five R-indices displayed significant associations with each other, with the most prominent correlations observed among R3, R4 and R5 (Pearson’s r values 0.39-0.46, **Figure 2c**).

### Association with demographics

All R-indices showed significant correlations with chronological age, with the strongest correlations observed between R5 and age (**Figure 2d**). In contrast, R1 and R2 showed relatively lower correlations. Additionally, tested through the Fisher r-to-z transformation, we observed small but significant differences between male and female groups in correlations between age and R1 (p=1.6*10^-4^), R3 (p=2.7*10^-5^), and R5 (p=7.6*10^-4^), suggesting potential influences from sex-related factors.

### Association with chronic diseases

To investigate the relationship between R-indices and chronic disease risk, UKBB individuals with a lifetime diagnosis of a chronic disease were grouped into 14 disease categories: MCI/dementia, stoke, multiple sclerosis, hypertensive diseases, diabetes, depression, bipolar disorder, schizophrenia, Parkinson’s disease, chronic obstructive pulmonary disease, osteoarthritis, chronic kidney disease (CKD), osteoporosis, and ischemic heart disease. An additional 2550 participants from 6 other studies were included to enrich the MCI/dementia cohort. 2971 UKBB participants without any of the 14 diseases were categorized as a healthy control (HC) group. To test differences in R-indices between the control and diseased groups, we conducted multiple linear regression analyses, adjusting for age and sex (**Figure 3a**).

**Figure 3.**
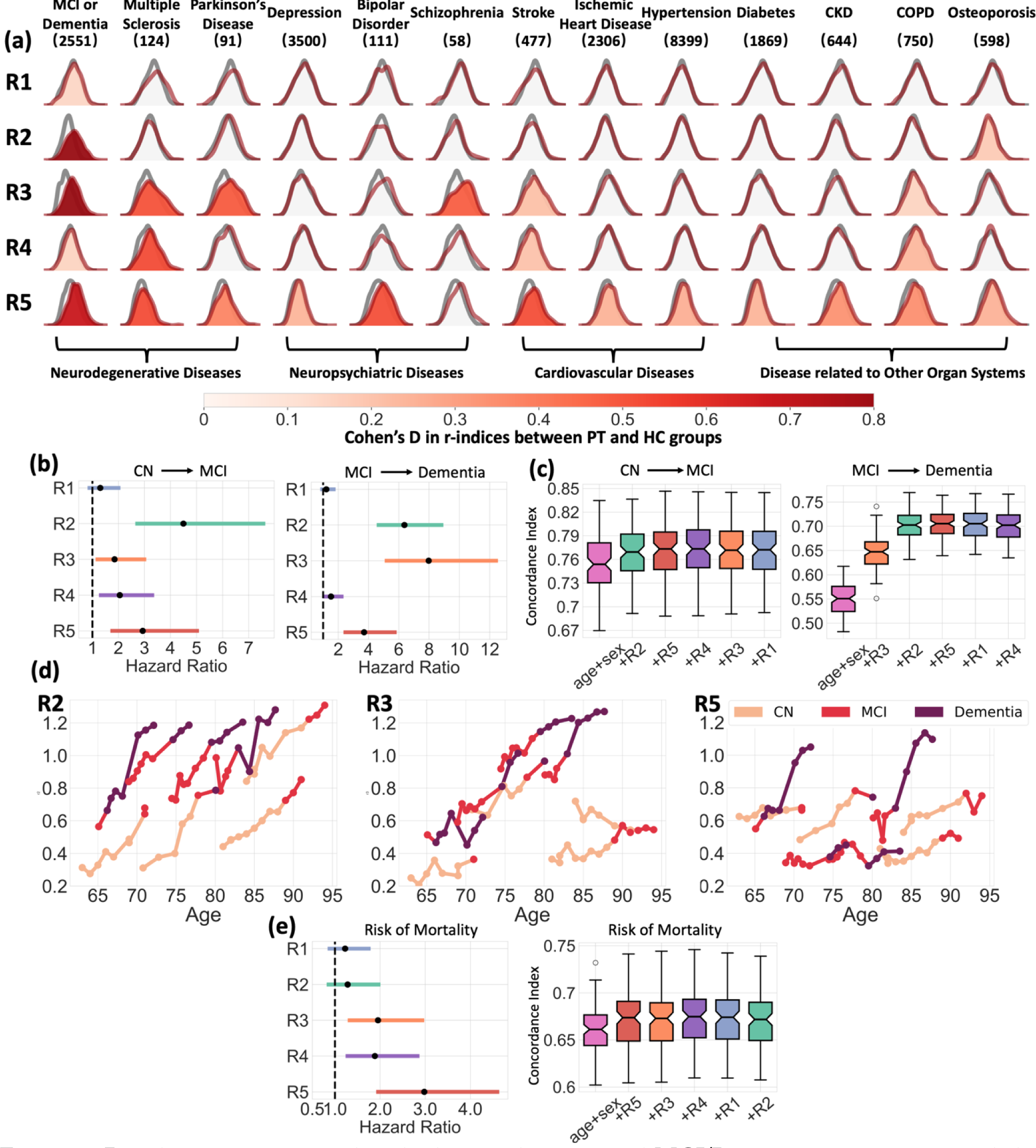
R-indices are associated with chronic diseases, and MCI/Dementia progression, and the risk of mortality. a. The distributions of R-indices are significantly different between the healthy control (HC) group and each patient (PT) group corresponding to one of the 13 chronic diseases, after adjusting for age and sex (p<6.6×10^-4^, Bonferroni-corrected). Warmer colors denote larger Cohen’s d (PT>HC). Distributions without color fill indicate no significant difference from the HC group (grey). **b** Values of the R2-5 indices exhibit associations with the risk of progression to MCI or dementia, as indicated by the corresponding hazard ratios. Cox proportional hazard models were used for testing associations, adjusting for age and sex. **c** The R-indices contribute to enhanced performance in predicting disease progression. Based on the significance of R-indices demonstrated in **(b),** we progressively incorporated R-indices as features one by one when fitting the Cox proportional hazard model on participants over 60 years old at baseline. For each combination of features, 100 iterations of 20% holdout cross-validation were performed to derive concordance indices. **d** The progression paths in R2, R3, and R5 of eight representative participants transitioning either from CN to MCI or from MCI to dementia. Different colors represent the distinct diagnoses. **e** The baseline R5 shows significant associations with the risk of mortality, with age and sex adjusted as covariates in Cox regression. Similar to **(c)**, R-indices were progressively included as features in cross-validation for mortality risk prediction among participants over 60 years old.

The MCI/dementia cohort exhibited significantly advanced R-indices along all five dimensions, with the most prominent effects observed in R2 (Cohen’s d (d)=0.869, p=4.1*10^-131^), R3 (d =0.849, p=1.6*10^-125^), and R5 (d=0.673, p=5.7*10^-81^). Patients with multiple sclerosis showed advanced aging along R3 (d=0.498, p=6.7*10^-8^), R4 (d=0.515, p=2.4*10^-8^), and R5 (d=0.513, p=2.8*10^-8^ ). Additionally, participants with schizophrenia (d=0.469, p=4.1*10^-4^ ) and Parkinson’s disease (d=0.481, p=9.6*10^-6^) demonstrated more severe aging effects along the R3 dimension. Furthermore, the R5 dimension was associated with neuropsychiatric diseases, as well as a group of chronic diseases related to other organ systems, including the respiratory, renal, metabolic, and cardiovascular systems. Detailed results between R-indices and all diseases can be found in **Supplementary Data 1**.

### Baseline R-indices predict disease progression and mortality

In addition to exploring baseline associations with chronic diseases, we examined the prognostic potential of R-indices for future progression from CN to MCI and from MCI to dementia using 2700 participants with longitudinal data from seven studies. The Cox proportional hazard model was utilized to test the associations between R-indices and the risk of progression, while adjusting for age and sex. R2 (p=2.7*10^-8^, HR (95% CI) = 4.50 (2.65, 7.64)), R4 (p=4.5*10^-3^, HR (95% CI) = 2.05 (1.25, 3.37)), and R5 (p=1.3*10^-4^, HR (95% CI) = 2.93 (1.69, 5.09)) were significantly associated with the risk of CN to MCI conversion, with R2 demonstrating the strongest prognostic indicator (**Figure 3b**). R2 (p=1.1*10^-26^, HR (95% CI) = 6.37 (4.54, 8.94)), R3 (p=2.8*10^-19^, HR (95% CI) = 7.97 (5.07, 12.54)), and R5 (p=2.2*10^-8^, HR (95% CI) = 3.70 (2.34, 5.86)) were significantly associated with the risk of progression from MCI to dementia, with R2 and R3 being the leading prognostic indices (**Figure 3b**). In cross-validation on participants over 60 years old, combining R2 (CI=0.768±0.031) and R5 (CI=0.773±0.033) as features in addition to age and sex (CI=0.755±0.033) improved the prediction for the risk of CN to MCI progression. Similarly, for predicting progression from MCI to dementia, the addition of R3 (CI=0.646±0.035), R2 (CI=0.704±0.030), and R5 (CI=0.705±0.030) significantly enhanced the predictive performance (CI=0.550 ± 0.034) (**Figure 3c**). Furthermore, in **Figure 3d**, we showcased the longitudinal progression paths of R2, R3, and R5 for eight representative participants who experienced disease conversions. In alignment with the baseline risk associations, these representative participants exhibited rapid increases in R2 before transitioning from CN to MCI, and rapid increases in R2 and R3 before progressing from MCI to dementia. Significant increases in R5 were also observed among some participants following their diagnoses of dementia.

Using similar approaches, we then examined the risk of mortality using UKBB participants (**Figure 3e**). R3 (p=1.8*10^-3^, HR (95% CI) = 1.95 (1.28, 2.98)), R4 (p=3.2*10^-3^, HR (95% CI) = 1.89 (1.24, 2.87)), and R5 (p=1.3*10^-6^, HR (95% CI) = 2.98 (1.91, 4.64)) showed significant associations with the risk of mortality, with R5 being the most significant prognostic indicator. Among participants over 60 years old, the combination of R5 (0.672 ± 0.029), R3, and R4 (0.674±0.029) as features, along with age and sex (CI = 0.661±0.027), improved the prediction for the risk of mortality, while the subsequent addition of other R-indices resulted in slightly decreasing predictive performances.

### Associations with clinical variables

We further explored associations between the five dimensions and cognition, as well as CSF/plasma biomarkers. Using partial correlations, we first tested associations of R-indices with four ADNI composite scores within the ADNI cohort (N=2214), along with four other cognitive scores among participants from multiple studies (N=6280-30444, **Figure 4a**). We observed consistent and significant associations among R2, R3, and R5 and all cognitive scores, although the pattern of associations differed between the cognitive scores. Specifically, R2 exhibited a particularly pronounced correlation with memory performance ( p = 0.462 , p=1.8*10^-117^, measured by ADNI-MEM ), while R3 demonstrated similar associations with executive function ( p = 0.349 , p=9.8*10^-65^ vs ADNI-EF) and memory ( p = 0.343, p=3.2*10^-62^ vs ADNI-MEM).

**Figure 4.**
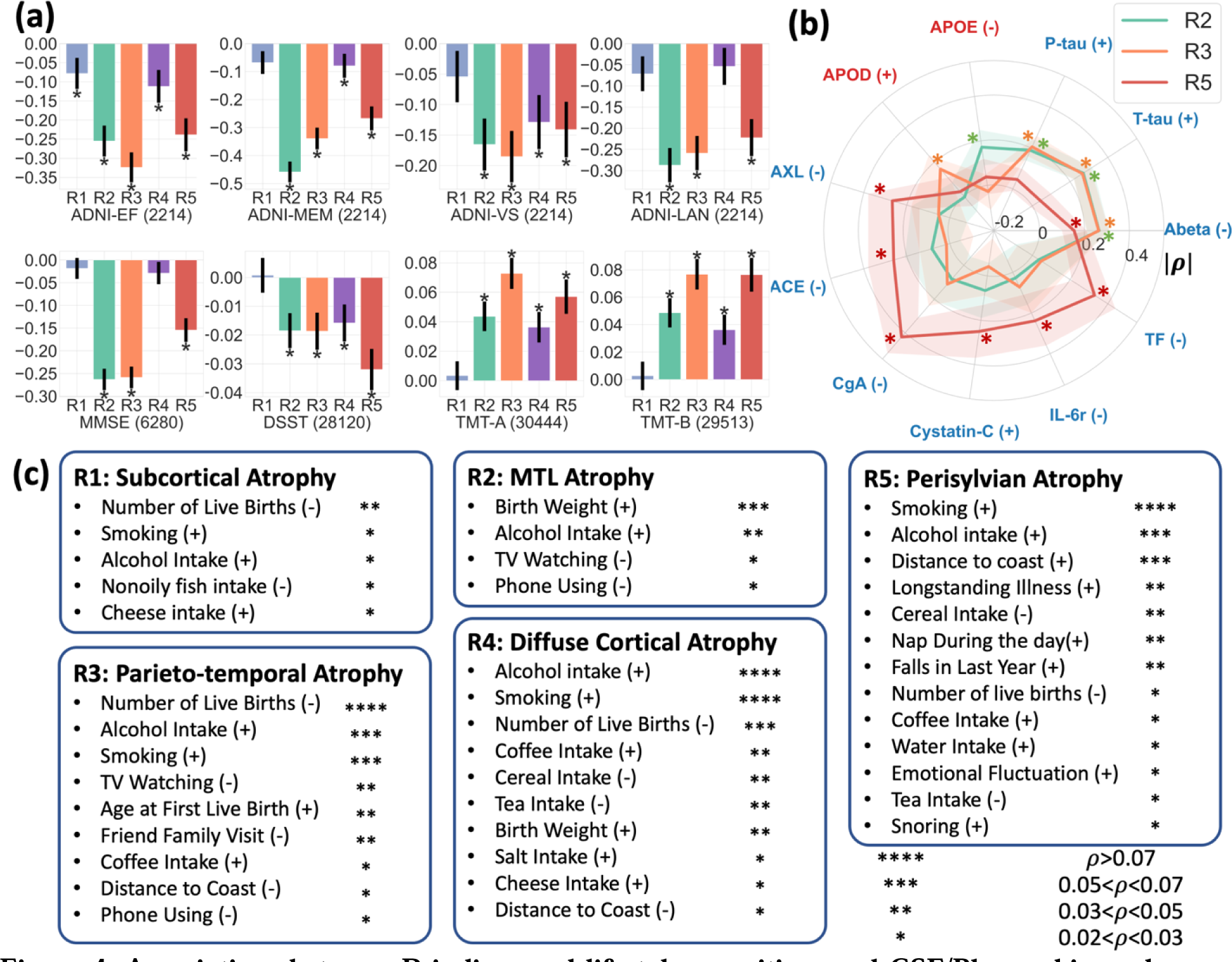
Associations between R-indices and lifestyle, cognition, and CSF/Plasma biomarkers. a. The five R-indices have distinct levels of association with different cognitive variables. Partial correlation was used for testing the associations between R-indices and cognitive scores, adjusting for age and sex. Additional site adjustments were performed for MMSE, DSST, TMT-A, and TMT-B to account for the utilization of multi-site data. Significantly associated R-indices are marked by * (p< 1.25X10^-3^ , Bonferroni-corrected). **b.** Among the R-indices, R2, R3, and R5 have significant associations (marked by *) with 11 CSF/plasma biomarkers obtained from the ADNI study (p<6X10^-4^). The CSF biomarkers are labeled in blue, and the plasma biomarkers are labeled in red. The radial graph presents the values and standard deviations of the correlation coefficients. For easier visualization, we invert the signs of negative coefficient (denoted by |p|) when making the plot. The “+” and “-“ signs alongside the biomarker names indicate positive and negative correlations. Due to the small sample sizes, the Benjamini-Hochberg procedure was used for FDR correction. **c.** The five R-indices show significant associations with a group of environmental/lifestyle factors and life events from the UKBB study (p< 8.7X10^-5^ , Bonferroni-corrected). Partial correlation adjusting for age and sex was used, as in (**a**). The number of “*” indicates correlation coefficients (legend). Positive and negative associations are denoted by “+” and “-” signs respectively, adjacent to the factor names.

Among the 232 CSF/plasma biomarkers collected in the ADNI study, we identified 15 significant associations between R-indices and biomarkers (**Figure 4v**). Among them, R2 and R3 revealed significant positive associations with CSF-pTau181 (p=1.6*10^-12^ vs R2; p=1.3*10^-14^ vs R3) and negative associations with CSF-Abeta42 (p=2.6*10^-23^ vs R2; p=4.4*10^-23^ vs R3), two hallmarks of Alzheimer’s disease^12^. Binary amyloid and tau positivity also showed significant associations with R2 (p=0.290, p=7.0*10^-28^ vs amyloid+; p ^-10^ vs tau+) and R3 (p ^-22^ vs amyloid+; p =0.173, p=1.4*10^-5^+.) R5 was associated with a group of other biomarkers, including chromogranin-A (p=1.3*10^-11^), tissue factor (p=2.6*10^-7^), AXL receptor tyrosine kinase (AXL) ( p=3.5*10^-5^ ), angiotensin-converting enzyme (ACE) (p=5.4*10^-5^), cystatin-C (p=1.1*10^-5^), and interleukin-6 receptor (IL-6r) (p=2.4*10^-4^), which potentially reflected the underlying hemostatic and inflammatory mechanisms^13-15^.

### Association with environmental and lifestyle factors

We examined the influence of lifestyle and environmental factors on variations along the five dimensions. Using partial correlations, we assessed the associations between the R-indices and 120 variables from the UKBB study, adjusting for age and sex (**Figure 4b**). In relation to all five atrophy dimensions, we found that alcohol intake has a statistically significant association with brain atrophy. Furthermore, smoking status primarily was associated with the expressions of R3 to R5. Notably, R4 and R5 were the two dimensions most associated with these two types of lifestyle factors. Moreover, the R3 dimension revealed significant associations pregnancy and social-recreational activities; the R4 dimension showed additional associations with various dietary habits; and the R5 dimension was exclusively associated with long-term illness, emotional factors, sleep, and environmental factors. Detailed correlations and p-values can be found in **Supplementary eData 2**.

### The five R-indices were associated with genetic variants

GWAS (**Method 10a**) identified 16, 17, 13, 9, and 18 genomic loci significantly associated with R1-5, respectively (**Figure 5a, Supplementary eFigure 3, and eData 5**). Among them, 38 loci were never associated with any clinical traits in the EMBL-EBI GWAS Catalog^16^, including 11, 7, 6, 5, and 11 loci for the five R-indices, respectively.

**Figure 5.**
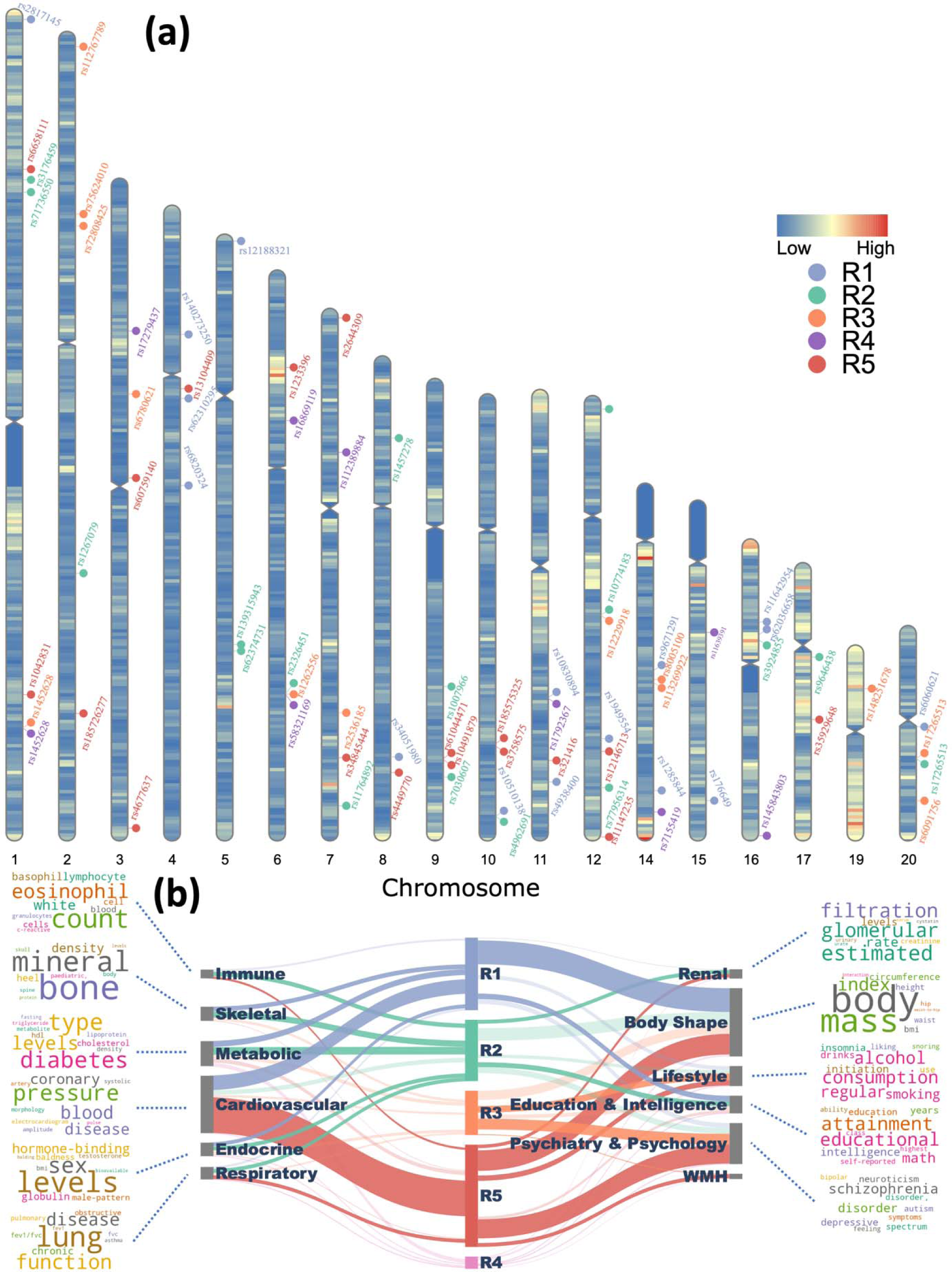
Five R-indices were associated with genomic loci that are novel or previously identified for other clinical traits. a. 73 genomic loci were associated with the five R-indices using a genome-wide P-value threshold [–log_10_(p) > 7.30]. For visualization purposes, we annotated the locus with the top lead SNP. **b** Phenome-wide associations of our identified genomic loci in the EMBL-EBI GWAS Catalog (query date: 2^nd^ July 2023 via FUMA version: v1.5.4). We examined the candidate and independent significant SNPs within each genomic locus and connected them to various clinical traits through a comprehensive query. The width of each connection represented the number of associations between the genomic loci revealed in our study and clinical traits in the literature. These traits were grouped into high-level categories, including different organ systems, psychiatric and psychological conditions, and lifestyle factors, body shape, etc. To enhance visual understanding of each category, we generated keyword cloud plots based on the frequency of clinical traits within each category. We excluded brain structure-related traits which were expected to have the highest number of associations with the SNPs we identified.

Using the GWAS catalog, we performed a phenome-wide association query (**Method 10b**) of the genetic variants previously identified in our GWAS. Specifically, genetic variants in our GWAS were previously associated with a total of 3895 clinical traits related to multiple organ systems and lifestyle factors. As expected, brain volume metrics and white matter microstructure were the most associated traits for genetic variants of all five R-indices. Additionally, the genomic loci associated with five R-indices were also enriched in many traits related to other organ systems (**Figure 5b**). For instance, R1 and R5 loci were enriched in traits related to body shape and cardiovascular system. R3 and R5 loci were linked to traits related to psychiatry and psychology, including schizophrenia, depression, worry feelings, etc. Also, R5 loci were specifically associated with WMH-related traits. Interestingly, through gene-set enrichment analyses (**Method 10c**), we found that the R1 dimension was enriched in the biological pathway of response to cortisol (p=1.1*10^-6^), which is related to the mediation of stress.

## Discussion

Human brain aging is affected heterogeneously by a complex interplay of genetic, lifestyle, and pathological factors. Our study dissects the heterogeneity of neurodegenerative changes related to aging, and reveals the predominant patterns of neurodegeneration, as well as their genetic, clinical, neuropathological, cognitive, and lifestyle correlates. Importantly, these analyses leveraged the extension of a recently developed deep representation learning method, Surreal-GAN, by adding a correlation structure in its latent space, which allowed us to better disentangle co-occurring and partially overlapping patterns of brain atrophy. Moreover, it utilized a large and diverse dataset of pooled and statistically harmonized MRI studies from older adults. Our analyses identified five reproducible dimensions of neurodegeneration: R1, subcortical atrophy; R2, MTL atrophy; R3, parieto-temporal atrophy; R4: diffuse cortical atrophy; and R5: perisylvian atrophy. Critically, our approach allows for evaluations of individualized levels of expression along these five dimensions, as quantified by respective R-indices, thereby offering additional tools for personalized patient management and clinical trial stratification.

We found correlations among the expressions of the five dimensions, indicating a co-expression of corresponding brain atrophy patterns at varying degrees. This underscores the interconnection and co-occurrence of underlying biological or neuropathological mechanisms, suggesting that the R-indices potentially measure the impact of multiple co-pathologies on the brain at the individual level. Notably, even though chronological age was not included in model training, the derived R-indices still exhibit significant positive associations with chronological age, highlighting Surreal-GAN’s ability to identify aging-related brain changes.

The dimensional system illuminates the intricate relationships between pathological factors and variations in brain aging. Conditions like multiple sclerosis show correlations with dimensions R3 to R5, potentially suggesting involvement of multiple cortical systems in a disease^17,18^ whose lesions can indeed intersect multiple brain networks. R3’s associations extend to schizophrenia and Parkinson’s disease, aligning with established cerebral connections^19–21^. Specifically, in schizophrenia, the medial temporal gyrus’s involvement is underscored by its role in auditory verbal hallucinations (AVH), a prominent symptom of this disorder^22^. R5 exhibits broad associations with various systemic diseases, including neuropsychiatric disorders and cardiovascular diseases, and immune health factors, which is consistent with its strongest associations with WMH volumes. These findings might be partially explained by the role of insular cortex in autonomic regulation^23–25^ and emotion processing^26,27^. Recent studies have increasingly recognized the role of the insula in depression and bipolar disorder specifically related to disordered interoceptive function^28^. White matter on the other hand has been shown to be primarily related to depression with a causal role in its etiology having postulated^29^. Furthermore, the co-occurrence of distinct diseases in the same dimensions can provide insights into their shared symptoms and the increased mutual risk. For example, schizophrenia, Parkinson’s disease, and dementia, all represented in the R3 dimension, exhibit common symptoms such as cognitive decline and hallucinations, with schizophrenia and Parkinson’s disease associated with a higher risk of dementia^30,31^.

Projecting patient cohorts with specific diseases onto our dimensional system offered insights into disease heterogeneity. Various chronic diseases, including MCI/Dementia, multiple sclerosis, and Parkinson’s disease, exhibited elevated expressions across multiple dimensions. Individualized differences along these associated dimensions likely mirror their distinct phenotypic and pathological variations. For instance, Alzheimer’s disease (AD)^32^, a prevalent neurodegenerative condition among the elderly, presents considerable heterogeneity^33–35^. Among the five dimensions, R2, R3, and R5 are strongly correlated with MCI/dementia and display differential associations with main the AD-typical characteristics, including cognitive decline and abnormal amyloid and tau deposition^12^. R2 displays stronger associations with memory impairment, while R3 is more closely associated with executive dysfunction. In contrast to R2 and R3, R5 reveals weaker associations with amyloid and tau, but stronger associations with various other CSF biomarkers, potentially linking this dimension to underlying inflammatory and hemostatic mechanisms. This resonates with R5’s broad associations with distinct co-pathologies, especially vascular pathology, that have an additive role in dementia along with AD pathology. Recognizing these distinctions enables us to cluster participants using these concise R-indices, thus allowing the creation of more homogeneous subgroups for clinical trial recruitment or personalized treatment strategies.

Brain changes occurring along disease-associated dimensions may manifest during preclinical stages. R2, R3, and R5 indicate the risk of clinical progression along the AD continuum. R2 and R3 stand out as key predictors for the clinical conversion from CN to MCI and MCI to dementia, respectively. These results are consistent with early involvement of the hippocampus and MTL in AD, with spreading of the pathology along pathways connected to it, especially posterior parietal regions, aligning with the typical order of brain region involvement in AD^36^. Concerning mortality risk, chronological age and male sex emerge as the strongest risk factors. Controlling for them, baseline R-indices, particularly R5, retain significant prognostic values. These findings further underscore not only the clinical applicability of R-indices in disease prognosis but also the significance of uncovering interventions targeting factors that are associated with these dimensions to mitigate risk.

Our analyses of lifestyle and environmental factors elucidated additional correlates of the observed variations, thus suggesting potential interventions targeted at specific dimensions. Smoking and alcohol consumption, two important risk factors across organ systems, are negatively associated with cortical atrophy, primarily mapping to the R4 and R5 dimensions, with lesser effects on R3. In addition, daily dietary habits correlate with R4 and R5, having either negative (tea, cereal) or positive (cheese, coffee, salt) relationships. Directly managing lifestyle or investigating underlying mechanisms might yield feasible interventions, although further studies are needed to understand causal relationships. Expression of any of the 5 intermediate phenotypes identified herein can serve as an indicator of active involvement of respective genetic and lifestyle risk factors, thereby prompting more aggressive patient management as well as recruitment to respective clinical trials.

Beyond daily life factors, certain life experiences also have associations with brain aging across dimensions. R1’s and R3’s associations with pregnancy-related factors partially explain their slightly larger deviation from the chronological age in females. Notably, childbirth has been shown to exert a long-term influence on women’s brain age in late life^37^, possibly contributing to a “young-looking” brain presumed to be partially related to fluctuations in hormonal or inflammatory mechanisms. Our study advances understanding by mapping these effects to specific dimensions of brain changes in addition to the overall brain age. In cognitively normal participants, R2 was negatively correlated with birth weight, resonating with previous findings of reduced hippocampal volumes and learning difficulties among preterm-born children with very low birth weights^38^.

Genetic variants influencing brain aging heterogeneity provide additional insights for intervention strategies, particularly in drug development. Our genetic analyses have revealed 73 SNPs associated with the five dimensions, including 38 SNPs without associated traits in the GWAS Catalog. The previously identified SNPs correlate with various clinical traits that validate our findings. For instance, R3 loci are linked to schizophrenia-related traits, while R5 loci show associations with cardiovascular and neuropsychiatric conditions, along with white matter hyperintensities. Additionally, the R1 dimension displays a correlation with the gene set linked to the response to cortisol. This suggests a potential stress-related impact on morphological changes in the striatum, the region connected to R1 and demonstrated to be highly influenced by stress^39,40^. The identification of these SNPs and associated genes might inform drug discovery or repurposing efforts for interventions targeting these dimensions.

The five dimensions derived herein are limited by the resolution and detail offered by MRI, namely patterns of regional atrophy of GM and WM, measures of small vessel ischemic disease, and expansion of CSF spaces. As such, they do not directly measure underlying neuropathologic processes that lead to these neurodegenerative changes. However, they do allow us to evaluate whether or not an individual with certain risk factors expresses patterns of brain changes that have been specifically linked to these risk factors. They can therefore offer opportunities for personalized patient management and clinical trial recruitment (**Figure 6**). In particular, the dimensions represented by R-indices enable the provision of more personalized therapeutic plans and lifestyle recommendations tailored to individual expression levels. Moreover, beyond facilitating targeted patient recruitment, the five R-indices also contribute to more optimized clinical trial design from several other perspectives. For instance, tracking changes in specific R-indices can significantly contribute to the assessment of trial effectiveness and boost its power^41^. Furthermore, a combination of R-indices and other clinical variables can be employed to establish stratified trial benchmarks due to their indication of individualized disease progression speed at the trial’s baseline.

**Figure 6.**
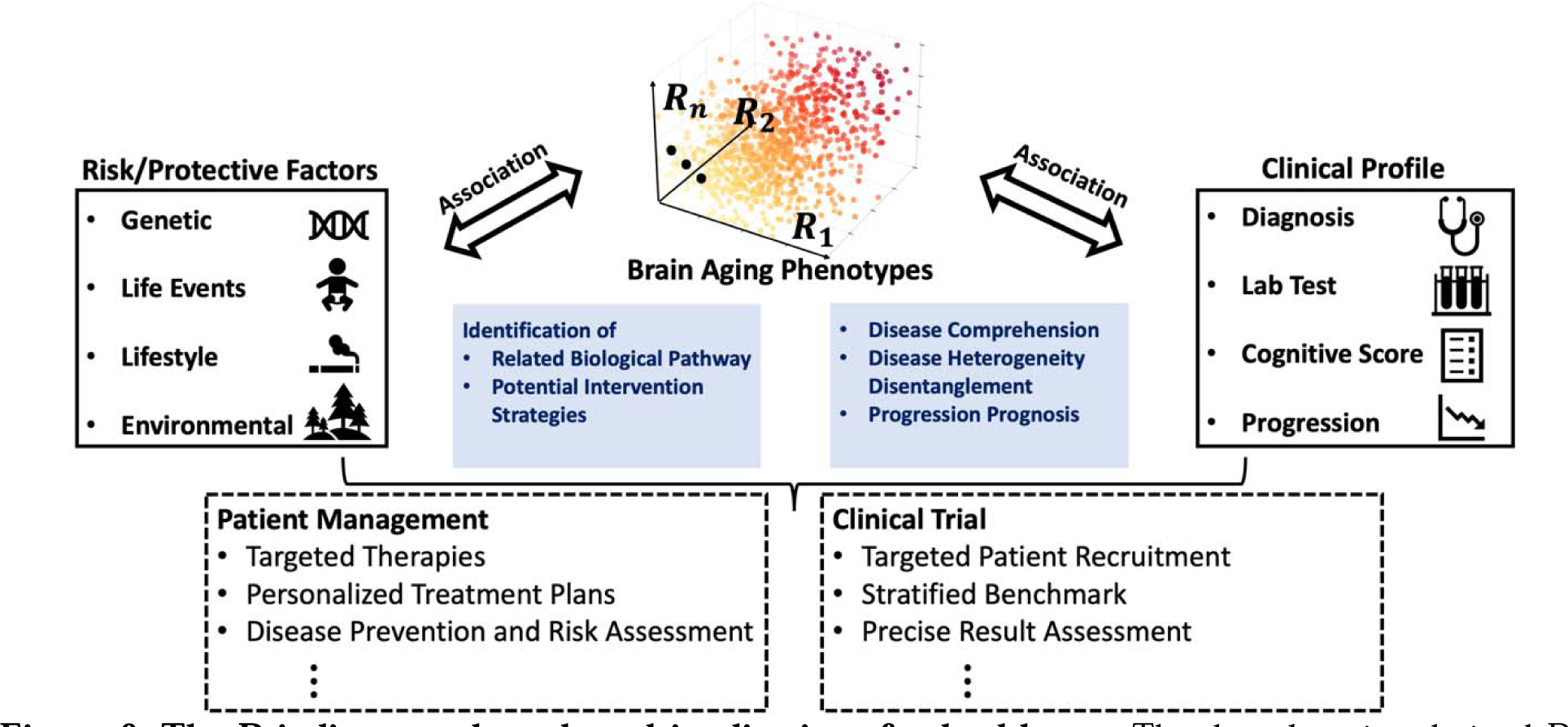
**The R-indices can have broad implications for healthcare**. The deep learning-derived R-indices are derived brain aging phenotypes that can serve as endophenotypes, or intermediate phenotypes, of diverse underlying neuropathologic processes that accompany aging. They also aid in understanding the risk and protective factors contributing to this heterogeneity. More importantly, these R-indices, combined with risk factors and clinical profiles, establish a concrete system for personalized patient management and targeted clinical trial recruitment designs.

In summary, this study characterized the neuroanatomical heterogeneity related to the aging process by offering a 5-dimensional representation system with five distinct brain signatures. As our consortium analyzes additional imaging data, including diffusion, functional, and images of tau and amyloid, it will continue to be extended and enriched. Currently, this dimensional system offers a means for dissecting the heterogeneity of neurodegeneration, as captured by sMRI, and to further understand its relationships to demographic, pathological, and lifestyle factors, as well as genetic variants. Moreover, it may contribute to personalized diagnostics and patient management, as well as to increased precision and effectiveness of clinical trials.

### Method 1. Surreal-GAN model

Surreal-GAN method^10^ is a weakly-supervised deep representation learning method for disentangling disease heterogeneity from neuroimaging data. Its key advantage lies in the ability to discern spatial and temporal (disease severity) variations solely from baseline data, thereby deriving low-dimensional R-indices that directly indicate the severity of distinct patterns of neuroanatomical changes. To capture phenotypic changes due to disease effects, Surreal-GAN learns multiple transformations from a reference (REF) group (e.g., pre-aging or health control) to a target (TAR) group (e.g., aging or patient)) through the generative adversarial network (GAN). Specifically, the method learns a function f to transform the REF data x into generated TAR data y′ = f(x, z), where z is a latent variable indicating the transformation directions. As is common in GAN-related methods^42^, an adversarial discriminator *D* is introduced to distinguish between real TAR data **y** and synthesized TAR data y′, thereby ensuring that the generated image data are indistinguishable from real patient data.

Beyond that, an inverse mapping, g, is introduced to re-estimate the latent variables z from the generated data y′ to ensure that the latent variables capture distinct and recognizable brain signatures. Multiple other regularizations were employed to further encourage the transformation function f to approximate the disease or aging effect, while boosting the positive association of different dimensions of the variable (z) with atrophy severity in distinct brain regions. The inverse function is utilized to derive the latent variables (referred to as R-indices) for real TAR data after the training process. More methodological details can be found in Yang et al^10^.

The original Surreal-GAN model^10^ derives independent R-indices, which limits its ability to characterize atrophy patterns driven by associated underlying pathologies. To address this limitation, we enhanced the Surreal-GAN method by parametrizing the covariance among latent dimensions using the Gaussian copula (**Supplementary eMethod 1.2**). This improvement enables the identification of associated R-indices, resulting in a significant boost in model’s performances and enhancing the method’s applicability (see **Supplementary eFigure 1** and **eResult 1** for details).

### Method 2. Study Population

The MRI (**Method 4**) and clinical (**Method 5, 6**) data used in this study were consolidated and harmonized by the Imaging-Based Coordinate System for Aging and Neurodegenerative Diseases (iSTAGING) study. The iSTAGING study comprises data acquired via various imaging protocols, scanners, data modalities, and pathologies, including more than 50,000 participants from more than 13 studies on 3 continents and encompassing a wide range of ages (22 to 90 years). Specifically, the current study used data from 52,319 participants from the Alzheimer’s Disease Neuroimaging Initiative (ADNI)^43^, the UK Biobank (UKBB)^44^, the Baltimore Longitudinal Study of Aging (BLSA)^45,46^, the Australian Imaging, Biomarker, and Lifestyle study of aging (AIBL)^47^, the Biomarkers of Cognitive Decline Among Normal Individuals in the Johns Hopkins (BIOCARD)^48^, the Open Access Series of Imaging Studies (OASIS)^49^, PENN, the Wisconsin Registry for Alzheimer’s Prevention (WRAP) studies^50^, the Coronary Artery Risk Development in Young Adults (CARDIA)^51^, Study of Health in Pomerania (SHIP)^52^, and the Women’s Health Initiative Memory Study (WHIMS)^53^. Among them, longitudinal data were available for 6576 participants. Detailed demographics and sample sizes from each study are detailed in **Table 1**. The aging participants analyzed in this study were predominantly of White ethnicity (N=44,539), constituting 95.1% of the total participants with reported race information. Race information was provided by all studies except SHIP. Detailed race distributions across different studies were provided in **Supplementary Data 6**, with race classification criteria introduced in **Supplementary Data 7.**

**Table 1.**
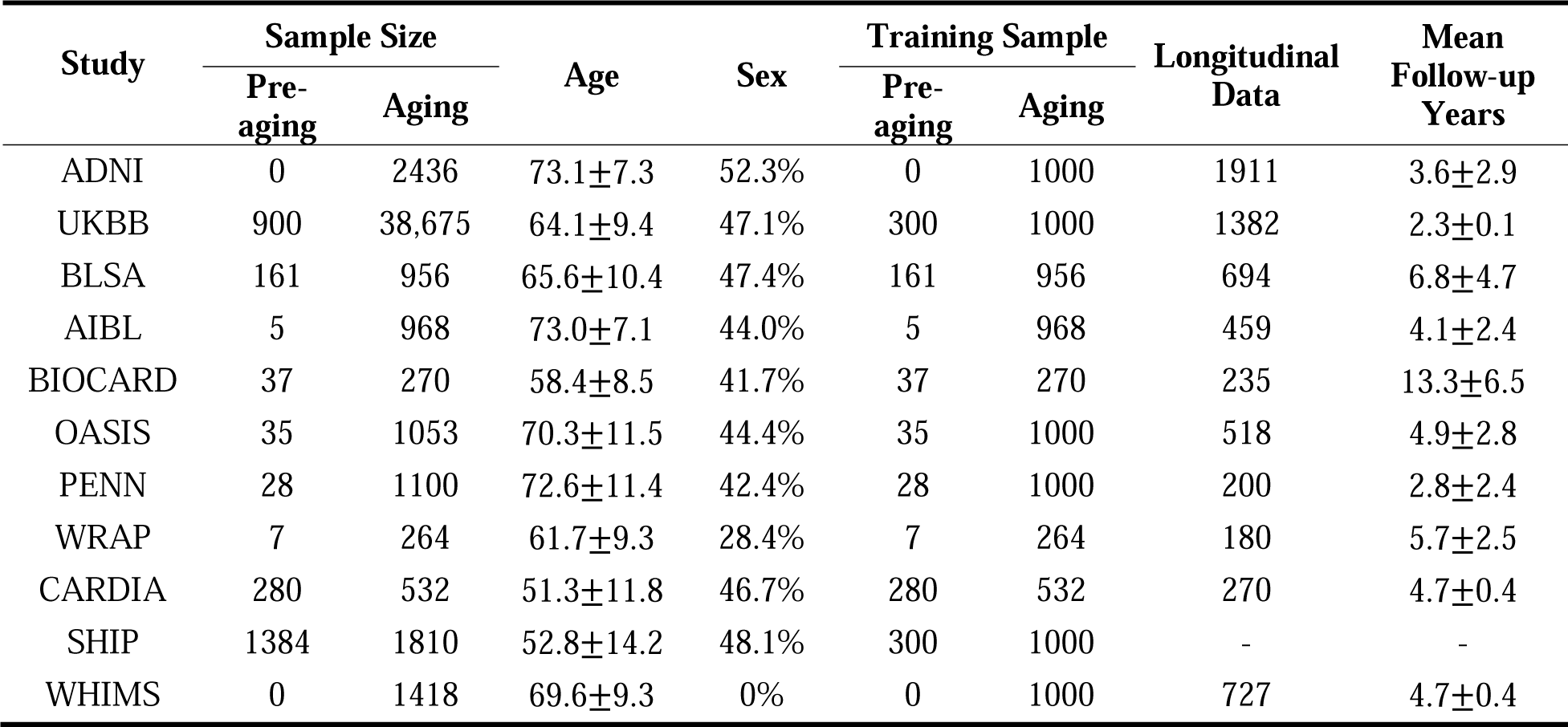
Participants and studies for model training and data analyses. For age and follow-up years, the mean and the standard deviation are reported. For sex, the percentage of males is presented. Longitudinal Data: number of participants with longitudinal data available.

| Study | Sample Size |  | Age | Sex | Training Sample |  | Longitudinal Data | Mean Follow-up Years |
| --- | --- | --- | --- | --- | --- | --- | --- | --- |
|  | Pre-aging | Aging |  |  | Pre-aging | Aging |  |  |
| ADNI | 0 | 2436 | 73.1±7.3 | 52.3% | 0 | 1000 | 1911 | 3.6±2.9 |
| UKBB | 900 | 38,675 | 64.1±9.4 | 47.1% | 300 | 1000 | 1382 | 2.3±0.1 |
| BLSA | 161 | 956 | 65.6±10.4 | 47.4% | 161 | 956 | 694 | 6.8±4.7 |
| AIBL | 5 | 968 | 73.0±7.1 | 44.0% | 5 | 968 | 459 | 4.1±2.4 |
| BIOCARD | 37 | 270 | 58.4±8.5 | 41.7% | 37 | 270 | 235 | 13.3±6.5 |
| OASIS | 35 | 1053 | 70.3±11.5 | 44.4% | 35 | 1000 | 518 | 4.9±2.8 |
| PENN | 28 | 1100 | 72.6±11.4 | 42.4% | 28 | 1000 | 200 | 2.8±2.4 |
| WRAP | 7 | 264 | 61.7±9.3 | 28.4% | 7 | 264 | 180 | 5.7±2.5 |
| CARDIA | 280 | 532 | 51.3±11.8 | 46.7% | 280 | 532 | 270 | 4.7±0.4 |
| SHIP | 1384 | 1810 | 52.8±14.2 | 48.1% | 300 | 1000 | - | - |
| WHIMS | 0 | 1418 | 69.6±9.3 | 0% | 0 | 1000 | 727 | 4.7±0.4 |

### Method 3. R-indices Derivation

For training the Surreal-GAN model, we included baseline data from 1150 participants below 50 years old as the REF group and that of 8992 participants over 50 years old as the TAR group including those with MCI or dementia. A maximum of 300 and 1000 individuals from each study were included in the REF and TAR groups, respectively, to avoid dominance from one study. All REF and TAR subjects were first residualized to rule out the sex and intracranial volume (ICV) effects estimated in the REF group using linear regression. Then, adjusted features were standardized with respect to the REF group. Without ground truth, we selected both the optimal number of dimensions, M (2-7), and hyper-parameters, γ and λ, by measuring agreements among repetitively trained models (**Supplementary eMethod 2.1**). For each of them, we repetitively trained the model 50 times and determined the optimal hyper-parameters leading to the highest agreements among the 50 models (M=5, γ=0.1, λ=0.8). Among the 50 corresponding models, the one having the highest mean pair-wise agreement with the other models was used to derive R-indices for all 49,482 elderly participants.

### Method 4. Image processing and harmonization

A fully automated pipeline was applied to process the T1-weighted MRIs. All MRIs were first corrected for intensity inhomogeneities.^54^A multi-atlas skull stripping algorithm was applied to remove extra-cranial material.^55^ Subsequently, 139 anatomical ROIs were identified in gray matter (GM, 119 ROIs) and white matter (WM, 20 ROIs) using a multiLJatlas label fusion method^56^. After merging symmetric ROIs from the left and right hemispheres, 72 ROI volumes were used as features for the Surreal-GAN model. Voxel-wise regional volumetric maps for GM and WM tissues (referred to as RAVENS)^57^, were computed by spatially aligning skull-stripped images to a single subject brain template using a registration method^58^. White matter hyperintensity (WMH) volumes were calculated through a deep-learning-based segmentation method^59^ built upon the U-Net architecture^60^, using inhomogeneity-corrected and co-registered FLAIR and T1-weighted images. Site-specific mean and variance were estimated with an extensively validated statistical harmonization method^61^ in the cognitively normal population and applied to the entire population while controlling for covariates.

### Method 5. Environmental/lifestyle factors and Clinical variables

From the UKBB study, we selected 120 variables that indicated individual differences in early life experience (e.g., birth weight, age at live births), lifestyle (e.g., smoking, alcohol consumption), social-recreation (e.g., frequency of friend-family visits, time spent watching television), psychological condition (e.g., mood status), local environmental exposures (e.g., coastal proximity, air pollution), and general health. Several variables were recoded for more convenient interpretation, as introduced in Tian et al^5^. The full list of selected variables was provided in **Supplementary eData 2**.

### Method 6. Cognitive, clinical, CSF, and plasma biomarkers

We included CSF and plasma biomarkers provided by ADNI, as well as cognitive test scores provided by nine studies. For ADNI, all measures were downloaded from the LONI website (http://adni.loni.ucla.edu). Detailed methods for CSF measurements of β- amyloid (Aβ) and phospho-tau (p-tau) are described in Hansson et al.^62^ Other CSF and plasma biomarkers were measured using the multiplex xMAP Luminex platform, with details described in “Biomarkers Consortium ADNI Plasma Targeted Proteomics Project – Data Primer” (available at http://adni.loni.ucla.edu). The ADNI study has previously validated several composite cognitive scores across several domains, including ADNI-MEM^63^, ADNI-EF^64^, and ADNI-LAN^65^. Four other cognitive scores were provided by different studies. (**Supplementary eData 4**)

### Method 7. Health outcomes

Based on the self-report (field 20002) and healthcare records (field IDs 41270 and 41271) from the UKBB study, we defined the patient group of 14 different chronic diseases, including MCI/dementia, stroke, multiple sclerosis, hypertensive diseases, diabetes, depression, bipolar disorder, schizophrenia, Parkinson’s disease, chronic obstructive pulmonary disease (COPD), osteoarthritis, chronic kidney disease (CKD), osteoporosis, and ischemic heart disease. **Supplementary eData 3** lists non-cancer illnesses, ICD9, and ICD10 codes related to each of the 14 disease categories. Additionally, the MCI/dementia category also includes 2550 participants from six other studies, including ADNI, AIBL, BIOCARD, BLSA, OASIS, and PENN. Individuals diagnosed with more than one disease category were assigned to multiple groups. 2971 UKBB participants without any of the 14 diseases were categorized as a healthy control (HC) group. Mortality data released on March 4, 2021, from UKBB (field 40000) were used for analyses of the risk of mortality. The dates of death were determined through data linkages to national death registries in the UK (documentation: https://biobank.ndph.ox.ac.uk/showcase/showcase/docs/DeathLinkage.pdf). 397 participants were confirmed dead after the acquisitions of brain MRIs at baseline.

### Method 8. Statistical Analyses on baseline variables

We first calculated the Pearson’s correlations between R-indices and chronological age, as well as among all R-indices. For testing associations between R-indices and all other baseline variables, we performed partial correlations with age and sex corrected as covariates. When comparing HC with each disease group in R-indices, we first adjusted for age and sex effects through multiple linear regression, and then calculated the Cohen’s d effect size between the two groups. For variables from multiple studies, the categorized study information was also incorporated as additional covariates. The FDR (false discovery rate) was controlled at 5% using the Benjamini-Hochberg procedure for CSF/Plasma biomarkers due to the small sample size. Bonferroni correction was used to control the family-wise error elsewhere.

Through the Python package Nilearn^66^, voxel-based morphometry (VBM) analyses were used for testing associations between voxel-wise tissue density and each r-index, adjusting for age, sex, ICV, and the remaining four R-indices. For VBM, The FDR (false discovery rate) was controlled at 0.1% using the Benjamini-Hochberg procedure.

### Method 9. Association with future risk of disease progression or mortality

To evaluate the association between each r-index and future progression from CN to MCI or from MCI to Dementia, we included 5807 participants with longitudinal diagnoses available, among whom 4777 were diagnosed as CN and 1030 were diagnosed as MCI at baseline. We employed a Cox proportional hazard model while adjusting for covariates such as age and sex to test the associations. The hazard ratio (HR) was calculated and reported as the effect size measure that indicates the influence of each r-index on the risk of disease progression. Further, to quantitatively assess prognostic performances with R-indices as features, we progressively added the most predictive r-indices to the Cox model to understand its optimal performance. The concordance index (CI) was utilized to quantify the performance of risk prediction in a 100 repetition of 20% hold-out CV. Using the same pipeline, we also evaluated the association between each r-index and the risk of mortality using all 38,675 participants from UKBB, with 397 confirmed dead after baseline assessments.

### Method 10. Genetic analyses

We used the imputed genotype data for all genetic analyses. Our quality check pipeline resulted in 32,829 participants with European ancestry and 8,469,833 SNPs. To summarize, we excluded individuals or genetic variants based on the following exclusion criteria: 1) related individuals (up to 2^nd^-degree) identified through family relationship reference^67^; 2) duplicated variants; 3) individuals whose self-acknowledged sex did not match genetically identified sex; 4) individuals with more than 3% of missing genotypes; 5) variants with minor allele frequency (MAF) of less than 1%; 6) variants with larger than 3% missing genotyping rate; iv) variants that failed the Hardy-Weinberg test at 1*10^-10^. To adjust for population stratification^68^, we derived the first 40 genetic principle components (PC) using the FlashPCA software^69^. Details of the genetic quality check protocol are described elsewhere^70,71^.

**(a) : GWAS**: For GWAS, we ran a linear regression using Plink^72^ for each r-index, controlling for confounders of age, sex, age-sex interaction, age-squared, age-squared-sex interaction, total intracranial volume, and the first 40 genetic principal components. We adopted the genome-wide P-value threshold ( 5*10^-8^ ) and annotated independent genetic signals considering linkage disequilibrium.

**(b)** : Phenome-wide association queries for the identified loci in GWAS Catalog:

We queried the significant independent SNPs within each locus in the EMBL-EBI GWAS Catalog (query date: 2^nd^ June 2023, via FUMA version: v1.5.4) to determine their previously identified associations with any other traits. We further grouped these traits into 11 categories for visualization and interpretation.

**(c)** : Gene-set Enrichment Analyses:

We conducted gene set enrichment analyses using gene sets from the MsigDB database (v6.2), Bonferroni correction was performed for all tested genes (*P*< 2.64*10^-6^ ) and gene sets (*P*<4.68*10^-6^). All other parameters were set by default in FUMA.

### Data Availability

The GWAS summary statistics generated in this study are provided in Supplementary Data files. Data used for this study were provided from several individual studies via data sharing agreements that did not include permission for us to further share the data. However, data from ADNI are available from the ADNI database (adni.loni.usc.edu) upon registration and compliance with the data usage agreement. Data from the UKBB are available upon request from the UKBB website (https://www.ukbiobank.ac.uk/). Data from the BLSA study are available upon request at https://www.blsa.nih.gov/how-apply. Data from the AIBL study are available upon request at https://aibl.org.au/. Data from the OASIS study are available upon request at https://www.oasis-brains.org/. Data requests for BIOCARD, PENN, WRAP, CARDIA, SHIP, and WHIMS datasets should be directed to M.S.A, D.A.W, S.C.J, L.J.L, K.W, and M.A.E respectively. Participant-level derived R-indices generated in this study will be provided within one month of receiving approval granted from respective studies.

### Code Availability

The software Surreal-GAN is available as a published PyPI package. Detailed information about software installation, usage, and license can be found at: https://pypi.org/project/SurrealGAN/0.1.1/. Custom code can be found at: https://github.com/zhijian-yang/SurrealGAN.

## Acknowledgments

Data used in this study are part of the iSTAGING study (representative: Christos Davatzikos), the Preclinical AD Consortium (Marilyn S. Albert), the Alzheimer’s Disease Neuroimaging Initiative (Michael W. Weiner), the Baltimore Longitudinal Study of Aging (Susan M. Resnick). The iSTAGING consortium is a multi-institutional effort funded by NIA by RF1 AG054409.

The Baltimore Longitudinal Study of Aging neuroimaging study is funded by the Intramural Research Program, National Institute on Aging, National Institutes of Health and by HHSN271201600059C. The BIOCARD study is in part supported by NIH grant U19-AG033655. SHIP is part of the Community Medicine Research net of the University of Greifswald, Germany, which is funded by the Federal Ministry of Education and Research (grants no. 01ZZ9603, 01ZZ0103, and 01ZZ0403), the Ministry of Cultural Affairs and the Social Ministry of the Federal State of Mecklenburg-West Pomerania. MRI scans in SHIP-START and SHIP-TREND have been supported by a joint grant from Siemens Healthineers, Erlangen, Germany and the Federal State of Mecklenburg-West Pomerania. The HANDLS study is supported by the

National institute on Aging Intramural Research Program, National Institutes of Health, Project AG 000513. The HABS-HD project is funded by grants from the National Institute on Aging (NIA): R01AG054073, R01AG058533. HABS-HD MPIs consists of Sid E O’Bryant, Kristine Yaffe, Arthur Toga, Robert Rissman, & Leigh Johnson; and the HABS-HD Investigators consists of Meredith Braskie, Kevin King, James R Hall, Melissa Petersen, Raymond Palmer, Robert Barber, Yonggang Shi, Fan Zhang, Rajesh Nandy, Roderick McColl, David Mason, Bradley Christian, Nicole Philips, Stephanie Large, Joe Lee, Badri Vardarajan, Monica Rivera Mindt, Amrita Cheema, Lisa Barnes, Mark Mapstone, Annie Cohen, Amy Kind, Ozioma Okonkwo, Raul Vintimilla, Zhengyang Zhou, Michael Donohue, Rema Raman, Matthew Borzage, Michelle Mielke, Beau Ances, Ganesh Babulal, Jorge Llibre-Guerra, Carl Hill and Rocky Vig. Other supporting grants include 5U01AG068057-02, 1U24AG074855-01. S.R.H and J.C.M are supported by multiple grants and contracts from NIH. A.A. was supported by grants 191026 and 206795 from the Swiss National Science Foundation. This research has been conducted using the UK Biobank Resource under Application Number 35148. Data used in the preparation of this article were in part obtained from the Alzheimer’s Disease Neuroimaging Initiative (ADNI) database (adni.loni.usc.edu). As such, the investigators within the ADNI contributed to the design and implementation of ADNI and/or provided data but did not participate in the analysis or writing of this report. A complete listing of ADNI investigators can be found at: http://adni.loni.usc.edu/wpcontent/uploads/how_to_apply/ADNI_Acknowledgement_List.pdf. ADNI is funded by the National Institute on Aging, the National Institute of Biomedical Imaging and Bioengineering, and through generous contributions from the following: AbbVie, Alzheimer’s Association; Alzheimer’s Drug Discovery Foundation; Araclon Biotech; BioClinica, Inc.; Biogen; Bristol-Myers Squibb Company; CereSpir, Inc.; Cogstate; Eisai Inc.; Elan Pharmaceuticals, Inc.; Eli Lilly and Company; EuroImmun; F. Hoffmann-La Roche Ltd and its affiliated company Genentech, Inc.; Fujirebio; GE Healthcare; IXICO Ltd.; Janssen Alzheimer Immunotherapy Research & Development, LLC.; Johnson & Johnson Pharmaceutical Research & Development LLC.; Lumosity; Lundbeck; Merck & Co., Inc.; Meso Scale Diagnostics, LLC.; NeuroRx Research; Neurotrack Technologies; Novartis Pharmaceuticals Corporation; Pfizer Inc.; Piramal Imaging; Servier; Takeda Pharmaceutical Company; and Transition Therapeutics. The Canadian Institutes of Health Research is providing funds to support ADNI clinical sites in

Canada. Private sector contributions are facilitated by the Foundation for the National Institutes of Health (www.fnih.org). The grantee organization is the Northern California Institute for Research and Education, and the study is coordinated by the Alzheimer’s Therapeutic Research Institute at the University of Southern California. ADNI data are disseminated by the Laboratory for Neuro Imaging at the University of Southern California. Mr. Yang had full access to all the data in the study. He took responsibility for the integrity of the data and the accuracy of the data analysis.

