## Supplementary Information for "Five dominant dimensions of brain aging are identified via deep learning: associations with clinical, lifestyle, and genetic measures"

**eMethod 1: Surreal-GAN: supplementary methodological details.**
**eMethod 1.1: Application of the inverse functions for deriving R-indices.**
**eMethod 1.2: Modification of the GAN loss in the Surreal-GAN model: motivation and derivation.**
**eMethod 2: Implementation details of the Surreal-GAN model.**
**eMethod 2.1: Hyperparameter selection.**
**eMethod 2.2: Training details.**
**eMethod 3: Agreement metric.**
**eMethod 4: Semi-synthetic experiments.**
**eMethod 5: Replication of the identified five dimensions.**

**eResult 1: The improved Surreal-GAN significantly outperforms the original version.**
**eResult 2: The five dimensions of brain aging are replicable with an independent training set.**

**eFigure 1: The improved Surreal-GAN significantly outperforms the original version.**
**eFigure 2: Surreal-GAN identifies five dimensions with replicable imaging signatures.**
**eFigure 3: Manhattan and QQ plots of baseline GWAS results for R1-R5 in the UKBB population.**

**eMethod 1: Surreal-GAN: supplementary methodological details.**

**eMethod 1.1: Application of the inverse function for deriving R-indices.**

In the stage of model inference following training, the inverse function, g, is applied to both the training and independent test data to derive their R-indices. Through GAN and regularizations during the training process, the learned transformation function f was considered a good approximation of the underlying pathological process, denoted by function h, such that $f(x,z) \approx h(x,\sigma(z))$, where $\sigma \in \Omega$ and $\Omega$ is a class of permutation functions that change the order of elements in the latent variables z. Since the orders of indices in the latent variables are unimportant and we can always reorder them to find the best matching, we rewrite the equation as $f(x,z) \approx h(x,z)$ without loss of generality. For any real TAR data, $\bar{y} = h(\bar{x}, r) \sim p_{tar}(y)$, we can estimate its ground truth R-indices, represented by r, through $g(\bar{y}) = g\big(h(\bar{x},r)\big) \approx g\big(f(\bar{x},r)\big) \approx r$.

**eMethod 1.2: Modification of the GAN loss in the Surreal-GAN model: motivation and derivation.**

In most GAN-based models[1,2], including Surreal-GAN[3], the latent variables or noise variables are sampled from fixed distributions (e.g., gaussian or uniform distribution) predefined before the training process. The fixed latent distributions mostly do not affect the models' performances in generating realistic data. However, they are problematic if we use an inverse function to re-estimate the latent variables of the data.

As introduced in **Supplementary eMethod 1.1**, we assume the inverse consistency, $g\big(f(x,z)\big) \approx z$, and equality in distributions, $p_{syn}(f(x,z)) \approx p_{tar}(y)$, after the training process. We can further derive that $p(g(y)) \approx p(g(f(x,z))) \approx p(z)$, where $p(g(y))$ is the distribution of the derived TAR participants' R-indices. Therefore, if we sample z from a standard multivariate uniform distribution as in the original Surreal-GAN, the covariance of the derived R-indices will be the identity matrix, which leads to bias when the ground-truth R-indices are correlated with each other.

In real applications, we can never attain the exact equality of distributions, $p_{syn}\big(f(x,z)\big) = p_{tar}(y)$. Thus, a slight deviation of the ground truth latent distribution from the standard multivariate uniform distribution does not significantly affect the models' performances. However, intensive correlations among the ground truth R-indices do affect the Surreal-GAN performance (**Supplementary eFigure1**) if we sample the latent variable from the uniform distribution.

To resolve this issue, we first construct a parametrized latent distribution $p_{\theta_z}(z)$ that can model intercorrelations among the dimensions. Gaussian copula, modeling dependence among variables, emerges as a logical and fitting choice:

$$p_{\theta_z}(z) = C^{Gauss}_{\theta_z}(z) = \Phi_{\theta_z}(\Phi^{-1}(z_1), \Phi^{-1}(z_2), \ldots, \Phi^{-1}(z_M))$$

where $\Phi^{-1}$ is the inverse cumulative distribution function of a standard normal and $\Phi_{\theta_z}$ is the joint cumulative distribution function of a multivariate normal distribution with the mean vector zero and the covariance matrix equal to a correlation matrix $\theta_z$. The resulting distribution, $p_{\theta_z}(z)$, has covariance equals $\theta_z$ and has the marginal distribution of each dimension to be uniform, U[0,1]. We further make changes to the GAN loss function so that z is implicitly sampled from the parametrized distribution, $p_{\theta_z}(z)$, which is optimized to approximate the ground truth latent distribution.

$$L_{GAN}(\theta_D, \theta_f, \theta_z) = E_{y \sim p_{tar}(y)}\big[\log\big(D(y)\big)\big] + E_{y' \sim p_{syn}(y')}\big[1 - \log\big(D(y')\big)\big] \quad (1)$$

$$= E_{z \sim p_{\theta_z}(z), y \sim p_{tar}(y)}\big[\log\big(D(y)\big)\big]$$

$$+ E_{z\sim p_{\theta_z}(z),x\sim p_{ref}(x)}\left[1-\log\left(D\big(f(x,z)\big)\right)\right] \quad (2)$$

$$= \int_D p_{\theta_z}(z)E_{y\sim p_{tar}(y)}\big[\log\big(D(y)\big)\big]d^M z$$

$$+ \int_D p_{\theta_z}(z)E_{x\sim p_{ref}(x)}\left[1-\log\left(D\big(f(x,z)\big)\right)\right]d^M z \quad (3)$$

$$= \int_D p_U(z)p_{\theta_z}(z)E_{y\sim p_{tar}(y)}\big[\log\big(D(y)\big)\big]d^M z$$

$$+ \int_D p_U(z)\, p_{\theta_z}(z)E_{x\sim p_{ref}(x)}\left[1-\log\left(D\big(f(x,z)\big)\right)\right]d^M z \quad (4)$$

$$= E_{y\sim p_{tar}(y),z\sim p_U(z)}\big[p_{\theta_z}(z)\log\big(D(y)\big)\big] + E_{z\sim p_U(z),x\sim p_{ref}(x)}\left[p_{\theta_z}(z)\left(1-\log\left(D\big(f(x,z)\big)\right)\right)\right] \quad (5)$$

We derive the updated GAN loss function (5) from the original objective function (1) presented in Goodfellow et al[1]. The updated loss function enables us to sample z from a uniform distribution, $p_U(z)$, but penalizes the losses with their probability under the distribution $p_{\theta_z}(z)$. In the training procedure, we sample one value of z for each x per batch instead of taking the expectation over all possible values of z. Therefore, to penalize both terms equally, the same modification is also applied to the first term of the equation (5), in which z was not originally included.

Additionally, to prevent $p_{\theta_z}$ from converging to an extreme distribution (e.g., two latent variables become completely correlated), we added a regularization term that controls the distance between $p_{\theta_z}(z)$ and $p_U(z)$. Therefore, the final modified GAN loss function equals:

$$L_{GAN}(\theta_D, \theta_f, \theta_{z_2}) = E_{y\sim p_{tar}(y),z\sim p_U(z)}\big[p_{\theta_z}(z)\log\big(D(y)\big)\big] + E_{z\sim p_U(z),x\sim p_{ref}(x)}\left[p_{\theta_z}(z)\left(1-\log\left(D\big(f(x,z)\big)\right)\right)\right] + \alpha D_{KL}\left(p_U(z)\middle|p_{\theta_z}(z)\right)$$

**eMethod 2: Implementation details of the Surreal-GAN model.**

**eMethod 2.1: Hyperparameter selection**

We set four robust hyperparameters, α=0.02, κ = 80, ζ = 80, μ = 500, and η = 6, to their default values, which minimally impact model performance[3]. To fine-tune the remaining hyperparameters, including λ, γ, and the number of patterns M, we employed an iterative approach. Models were repetitively trained, with each parameter configuration undergoing 50 runs. The consensus (**eMethod 3**) among the 50 outcomes guided the selection of optimal hyperparameters. This consensus-based strategy is underpinned by the notable correlation between agreement among these iteratively trained models and their inherent representational accuracy[3]. For optimal training epoch selection, the model underwent $1500000*\frac{\text{batch size}}{\text{TAR sample size}}$ epochs, with checkpoints saved every $45000*\frac{\text{batch size}}{\text{TAR sample size}}$ epochs when reconstruction loss and monotonicity loss were below $3*10^{-3}$ and $6*10^{-4}$, respectively. Consensus among repeatedly trained models once again guided the determination of the optimal epoch.

**eMethod 2.2: Training details**

Regarding optimization procedure, ADAM optimizer was used with a learning rate (lr) $1.6*10^{-4}$ for Discriminator, $8*10^{-4}$ for transformation function f and inverse function g, and $2.7*10^{-5}$ for the latent distribution, $p_{\theta_z}(z)$. $\beta_1$ and $\beta_2$ are set to be 0.5 and 0.999, respectively. Moreover, for all experiments, the batch size was set to 300. We parametrize the latent distribution (i.e., the Gaussian copula) using the lower-triangular factor of the correlation matrix given its special properties. Other model structures and training algorithms were the same to the original Surreal-GAN[3].

**eMethod 3: Agreement metric**

For hyper-parameter selection and evaluating result reproducibility, we constructed a metric designed to quantify the level of agreement between two sets of R-indices, $r^1$ and $r^2$. To assess their concordance, we computed two distinct correlations, as outlined below:

1. **Dimension-correlation** is defined as the average of M Pearson's correlations for all dimensions: $\frac{1}{M}(\sum_{i=1}^{M}\rho(r_i^1, r_i^2))$
2. **Difference-correlation** is defined as the average of M(M-1)/2 Pearson's correlations for all pairs of dimensions: $\frac{2}{M(M-1)}(\sum_{i=1}^{M}\sum_{j=i+1}^{M}\rho(r_i^1 - r_j^1, r_i^2 - r_j^2))$

For the derivation of both values, we attempted varying permutations of the second set of indices to identify the optimal alignment. The resulting correlations, referred to as "Rindices-Correlation", represent the means of these two measurements and serve as a quantitative indicator of the agreements between $r^1$ and $r^2$.

**eMethod 4: Semi-synthetic experiments**

Semi-synthetic data construction: To assess the improved Surreal-GAN's ability to capture associations among underlying pathologies, we compared two models using semi-synthetic datasets with simulated ground-truth disease patterns. We followed the mild atrophy dataset construction approach from the Surreal-GAN paper, partitioning 1392 cognitively normal subjects (age < 70 years) into a REF group (492 subjects) and a Pseudo-TAR group (900 subjects). For each participant in the Pseudo-TAR group (denoted as the $i_{th}$ subject), a three-dimensional pattern severity vector ($s_i$) was sampled from a multivariate uniform distribution ($s_i \sim U[0,1]$), representing the ground truth. We introduced three atrophy patterns to the 900 Pseudo-TAR subjects based on the sampled severity vectors, each involving distinct regions. Unlike the Surreal-GAN paper, we simulated different levels of correlations among three dimensions: **1. Singular Correlation**; correlations of 0.2, 0.4, 0.6 were only simulated between two dimensions, respectively. **2. Comprehensive Correlation**; correlations of 0.2, 0.4, 0.6 were simulated across all three dimensions. Both the improved and original Surreal-GAN models were repetitively trained 50 times on each semi-synthetic dataset with different levels of correlation. Rindices-Correlations were calculated between the 50 derived R-indices and the ground truth, $s$, to quantify their respective performances.

**eMethod 5: Replication of the identified five dimensions.**

To test the reproducibility of the identified five dimensions. We retrained the model using a completely independent training set and performed comparisons. Specifically, we resampled 1000 pre-aging and 4818 aging participants not included in the original training sets. A maximum of 500 individuals from each study were included in the REF group (i.e., pre-aging group), while up to 2000 individuals were allocated to the TAR group (i.e., aging group). The same procedure introduced in **Method 3** was used to derive R-indices for both the replication training set and all elderly participants. Voxel-based morphometry analyses examined brain change patterns associated with each dimension in the independent population. We further quantified reproducibility by calculating the Rindices-Correlation between reproduced and original R-indices.

**eResult 1: The improved Surreal-GAN significantly outperforms the original version.**
As revealed in **Supplementary eFigure 1**, a noticeable decline in the performance of the original Surreal-GAN model becomes evident as positive or negative correlations among the ground-truth dimensions increase. The performance degradation is even more pronounced when correlations are introduced among all three dimensions. Conversely, the improved Surreal-GAN exhibited consistent performance across datasets characterized by varying levels of simulated correlations. This robustness underscores its ability to effectively handle associations among underlying pathologies.

**eResult 2: The five dimensions of brain aging are replicable with an independent training set.**
As shown in **Supplementary eFigure 2**, the replication model, trained with an independent dataset, successfully identifies five dimensions associated with consistently reproducible brain changes. This is supported by a quantitative analysis, where the replication R-indices exhibit a high Rindices-Correlation of 0.752 with the original R-indices, further underscoring their high concordance.

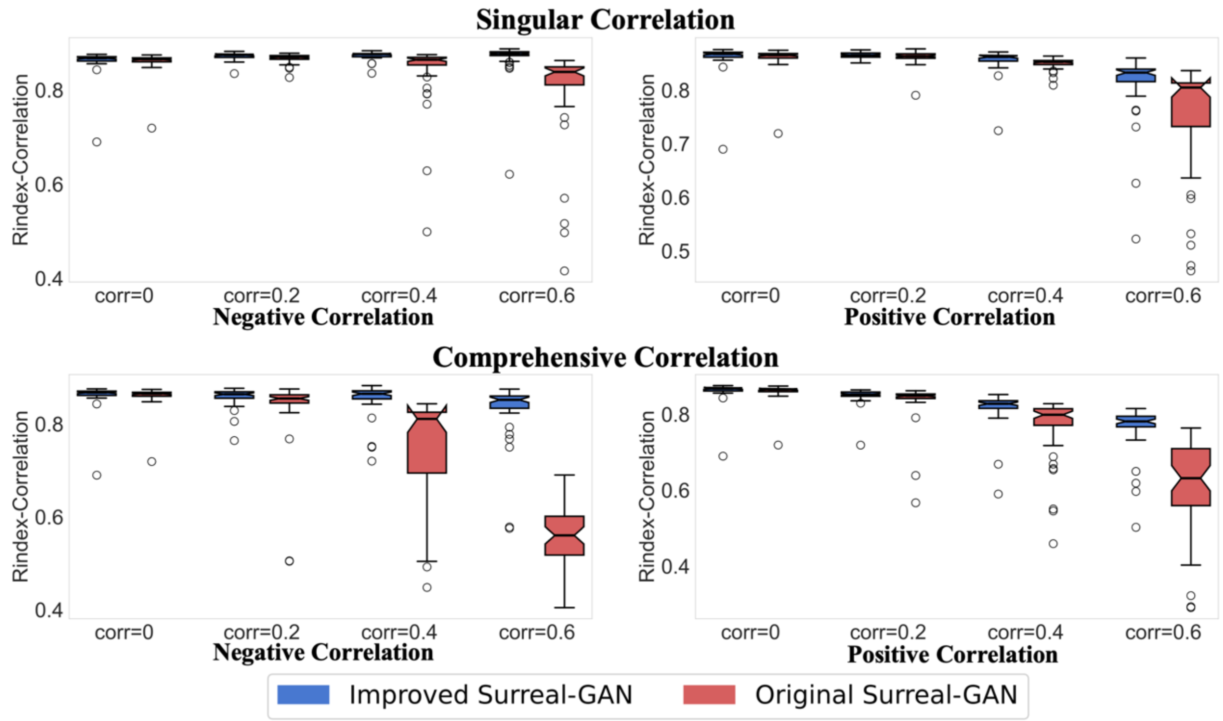


**Supplementary eFigure1. The improved Surreal-GAN significantly outperforms the original version.** With increased correlations simulated among the ground truth R-indices, the original Surreal-GAN shows dramatically decreased model performances. In contrast, the improved one demonstrates robustness, proving its capability in capturing the correlations among the ground-truth brain change patterns.

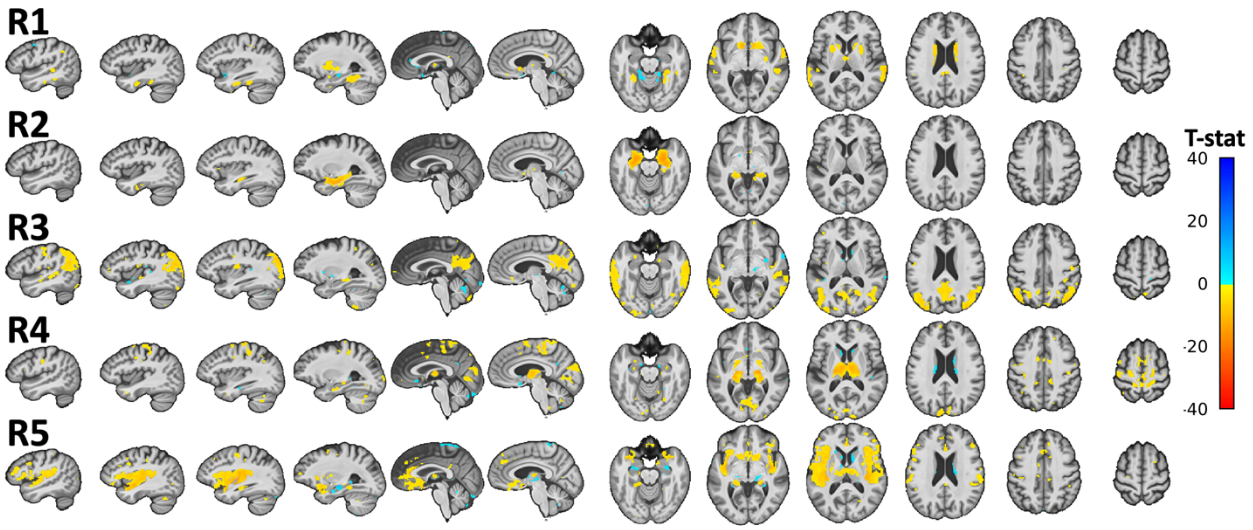


**Supplementary eFigure2. Surreal-GAN identifies five dimensions with replicable imaging signatures.** Voxel-wise t-tests were performed for the five dimensions rederived using an independent replication training set, while adjusting for age, sex, intracranial volume (ICV), and the remaining four R-indices. False discovery rate (FDR) correction was performed to adjust multiple comparisons with a p-value threshold of 0.001. The identified brain atrophy patterns generally replicate those revealed in **Figure 1a**.

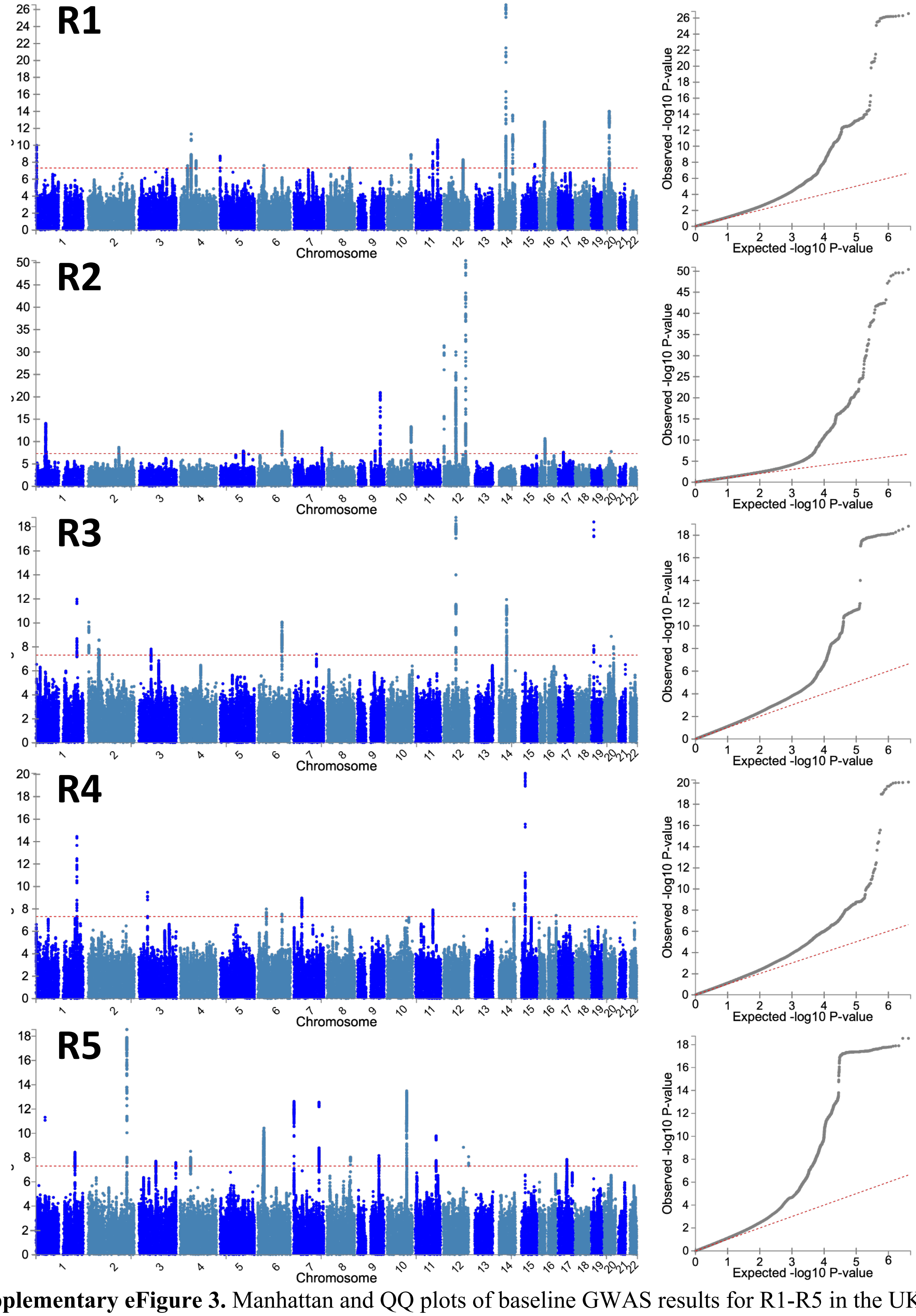


**Supplementary eFigure 3.** Manhattan and QQ plots of baseline GWAS results for R1-R5 in the UKBB population.

1 Goodfellow, I. *et al.* Generative Adversarial Networks. *Advances in Neural Information Processing Systems* **3** (2014). https://doi.org:10.1145/3422622
2 Chen, X. *et al. InfoGAN: Interpretable Representation Learning by Information Maximizing Generative Adversarial Nets*. (2016).
3 Yang, Z., Wen, J. & Davatzikos, C. Surreal-GAN:Semi-Supervised Representation Learning via GAN for uncovering heterogeneous disease-related imaging patterns. *International Conference on Learning Representations (ICLR)* (2022).
